# Prevalence of Lung Cancer in Colombia and a New Diagnostic Algorithm Using Health Administrative Databases: A Real-World Evidence Study

**DOI:** 10.1101/2022.05.16.22275161

**Authors:** Javier Amaya-Nieto, Gabriel F. Torres, Giancarlo Buitrago

**Author notes:** **Corresponding author:** Javier Amaya, Instituto de Investigaciones Clínicas, Universidad Nacional de Colombia, Calle 44 No. 45 - 67, Bogota, DC, Colombia. These authors contributed equally to this work.

## Abstract

Reliable, timely and detailed information on lung cancer prevalence, mortality and costs from middle-income countries is essential to policy design. Thus, we aimed to develop an electronic algorithm to identify lung cancer prevalent patients in Colombia by using administrative claims databases, as well as to estimate prevalence rates by age, sex and geographic region. We performed a cross-sectional study based on national claim databases in Colombia (*Base de datos de suficiencia de la Unidad de Pago por Capitación* and *Base de Datos Única de Afiliados*) to identify lung cancer prevalent patients in 2017. Several algorithms based on the presence or absence of oncological procedures (chemotherapy, radiotherapy and surgery) and a minimum number of months that each individual had lung cancer ICD-10 codes were developed. After testing 16 algorithms, those with the closest prevalence rates to those rates reported by aggregated official sources (Global Cancer Observatory and *Cuenta de Alto Costo*) were selected. We estimated prevalence rates by age, sex and geographic region. Two algorithms were selected: i) one algorithm that was defined as the presence of ICD-10 codes for 4 months or more (the sensitive algorithm); and ii) one algorithm that was defined by adding the presence of at least one oncological procedure (the specific algorithm). The estimated prevalence rates per 100,000 inhabitants were 15.3 and 9.7 for the sensitive and specific algorithms, respectively. These rates were higher in men (9.9 per 100,000), over 65-years-old (37.1 per 100,000) who lived in Central and Bogota regions (14.7 and 10.9 per 100,000, respectively). Selected algorithms showed similar aggregated prevalence estimations to those rates reported by official sources and allowed us to estimate prevalence rates in specific aging, regional and gender groups for Colombia by using national claims databases. These findings could be useful to identify clinical and economical outcomes related to lung cancer patients by using national individual-level databases.

## Introduction

Lung cancer has progressed to be one of the most frequent and deadly variations of cancer worldwide, even in low-middle-income countries (LMICs) [1]. Despite advances in diagnosing and treating this disease during the last decade, lung cancer prevalence and mortality have increased. According to the last report of The Global Cancer Observatory, this type of cancer represents the leading cause of mortality in cancer, with 23 deaths per 100,000 being reported globally [2,3]. Heterogeneity in prevalence trends within nations has been described, especially for LMIC countries showing lower incidence and prevalence rates than developed countries, which is a fact that has become a source of concern when considering that approximately 80% of all smokers live in LMIC [4]. Health outcomes are worse in Latin American countries than in other regions because of their fragmented and underbudgeted health systems, which presents an additional challenge in the process of reducing the incidence, prevalence and mortality rates of lung cancer [5]. For optimal cancer care, LMICs must demonstrate significant prevention and early diagnosis strategies, as well as high quality measures, in medication, surgery and radiation delivery backed by patient support. However, health systems in LMICs are precarious, and most people with this condition are diagnosed in late stages if their cancer occurs, thus causing higher mortality rates [6].

Lung cancer in Colombia has an important impact, as it represents the fourth place in mortality among men and the fifth place among women [7]. Furthermore, a demographic change in the population has been described, which indicates an increase in individuals aged over 60-years-old [8]. It has also been described that lung cancer risk increases over time, especially in elderly people over 65-years-old [9,10]. All of these combined factors can exacerbate some specific situations, such as increased prevalence rates in some regions of the country due to the high prevalence of smokers; additionally, mining activities have aroused interest over this specific problem [11,12].

The Colombian health system covers 99% of Colombian citizens [13]. It has a centralized fund controlled by the government that gathers payroll taxes and contributes an additional amount of money to ensure health care for the population, including the poorest individuals of this population. At the level of the insurers, there are public and private health institutions (known as *Empresas Promotoras de Salud* [*EPS*]) that act as intermediate payers between the centralized fund and health service providers (known as *Instituciones Prestadoras de Salud* [*IPS*]). These IPSs compete to be included in each EPS service network. Moreover, the insured population can be classified into a group that contributes to the health system via payroll taxes (known as the contributory regime) and a group funded by the national government (known as the subsidized regime) [14]. This distribution is important because it shows a degree of inequity and it also represents differences in prevalence estimation, health spending and clinical outcomes [15–18]. Furthermore, the benefit package of the health system is the same for both regimes and includes a comprehensive list of health technologies for the diagnosis and treatment of lung cancer.

This study aimed to develop an electronic algorithm to identify lung cancer-prevalent patients over 20-years-old in Colombia by using official databases, as well as to estimate prevalence rates of lung cancer by age, sex and geographic region. This information could provide valuable information to Colombian policy-makers, clinicians and the world to improve knowledge about lung cancer in LMICs.

## Methods

### Ethics and approval

This research was evaluated and approved by the ethics committee of the Universidad Nacional de Colombia as required by the national law [19].

### Design and population

We performed a cross-sectional study based on national claims databases in Colombia (*Base de datos de suficiencia de la Unidad de Pago por Capitación* [UPC] and *Base de Datos Única de Afiliados* [BDUA]) to identify lung cancer-prevalent patients in 2017. These databases contain information on the population that was registered yearly from 2015 to 2017; additionally, these databases provide information used for this research under academic agreements between the Ministry of Health (MoH) and *Universidad Nacional de Colombia*. All of the populations over 20-years-old were evaluated to identify individuals with lung cancer.

### Database characteristics

Several sources of information were used for this study. The first utilized source included the databases used by the MoH to calculate the sufficiency of the UPC that represents the amount of money that the government pays to IPS for each enrollee per year. It contained individual-level information about all of the health services that were used by each person in the system reported yearly by each IPS. Data provided in this database include the location of each service that was used, the date of utilization, the type of health service, the diagnostic code (ICD-10) associated with each health service, the cost of the service, the health provider and the municipality. This information was available at an individual level and provided for the 10 largest insurers in the contributory regime that represent 88% of the contributory regimes nationwide. Information registered in the UPC databases was validated through a validation process designed by MoH that involves correcting inconsistencies found in the data and the use of double validation with other databases used by the government. Information for the subsidized regime for Bogotá was also available by using the same information from the largest insurer in the city. The BDUA database includes sociodemographic and insurer (i.e., regime) information of all of the enrollees in Colombia.

Other secondary, aggregated and publicly available data sources were used. This information was collected from the Global Observatory of Cancer (GLOBOCAN) [20], National Cancer Institute (NCI)[21] in Colombia and *Cuenta de Alto Costo (CAC)* [22]. These data were used to verify the aggregated prevalence rates that were calculated in this study after having estimated differentiated prevalence rates over different age, region and sex groups.

### Algorithms of case identification

To identify individuals over 20-years-old with lung cancer diagnoses, this study used 16 different electronic algorithms. Some of the algorithms were based on identifying any ICD-10 codes for tumors of the trachea, bronchi or lungs with malignant behavior (for example, roots C33 and C34) during a minimum number of months and at least one oncological procedure in the same individual in the previous three years (2017, 2016 and 2015). We defined these algorithms as being specific. The second type of algorithm was known as the sensitive algorithm by our team and aimed to identify patients by solely using the ICD-10 codes for tumors of the trachea, bronchi or lungs with malignant behavior during a minimum period of time. Specific and sensitive algorithms were compared, and two of them were chosen by selecting the closest prevalence rates to those rates reported by aggregated official sources (GLOBOCAN, NCI and CAC). The number of patients with lung cancer was determined for different strata of age, geographical region and sex.

### Prevalence method description

For global prevalence, the following equation summarizes the method that was used in this paper:

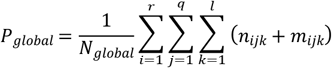

where ***P***_*global*_ is the global prevalence for Colombia in 2017; *N*_*global*_ is the total number of patients affiliated with the ten largest IPSs in the contributory regime, as reported by the BDUA, and the largest EPS from the subsidized regime by June 2017; *n*_*ijk*_ is the number of prevalent cases identified for each region *i*, age group *j* and sex *k* in the contributory regime; and *m*_*ijk*_ is the number of prevalent cases identified for each region *i*, age group *j* and sex *k* in the subsidized regime.

## Results

After testing 16 algorithms, two electronic algorithms were selected, as they had the closest prevalence rates to those rates reported by aggregated official sources (GLOBOCAN, NCI and CAC), as shown in Figure 1.

**Figure 1.**
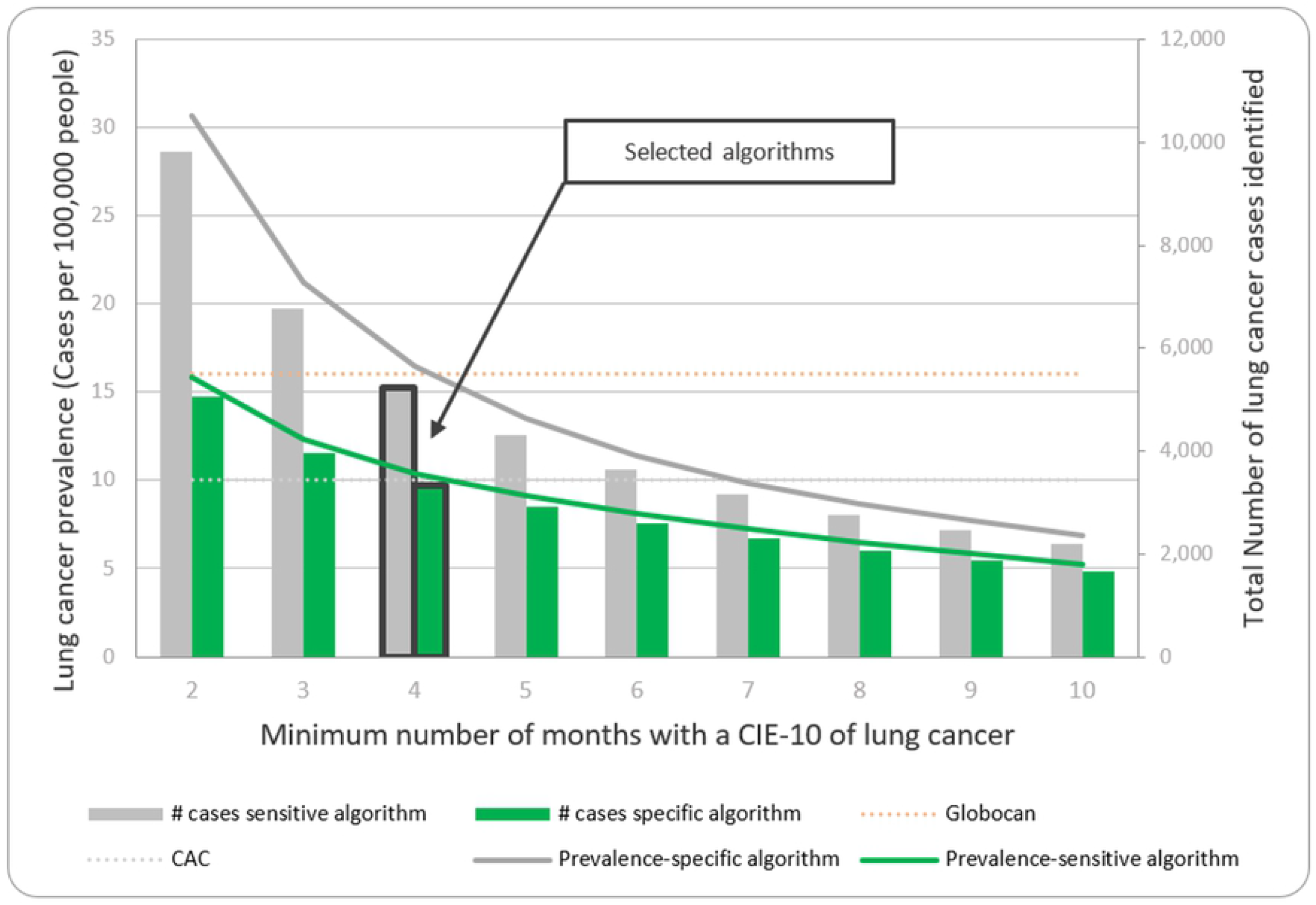
Total number of cases identified by two electronic algorithms developed for the study in comparison to aggregated official sources.

The first selected algorithm was defined as the presence of ICD-10 lung cancer codes for four months or more (the sensitive algorithm), and the second selected algorithm was defined as the same criteria from the first algorithm and the presence of at least one oncological procedure (the specific algorithm). Comparisons between the results that were obtained in this study and prevalence rates calculated from aggregated data prevalence rates are shown in Table 1.

**Table 1.**
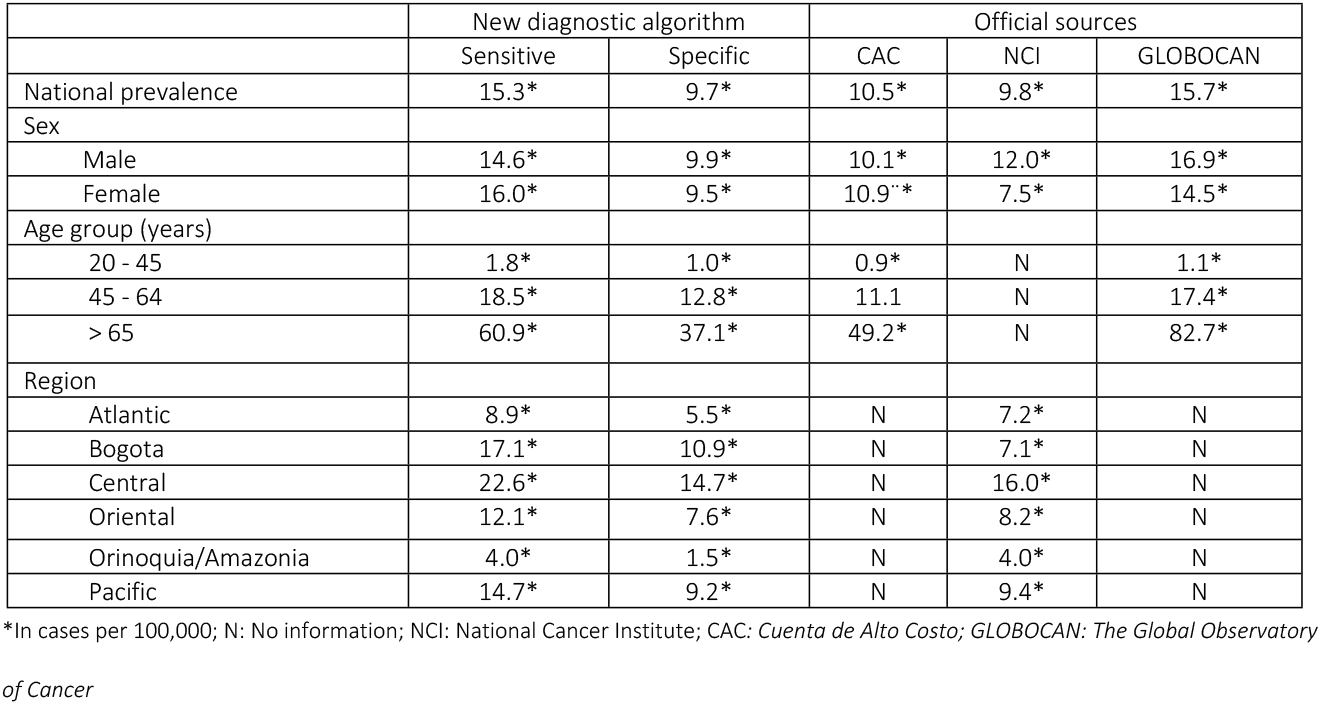
Comparison between the prevalence rates identified in this study versus official aggregated data sources

By using previously selected algorithms, the identified global prevalence rates were 15.3 and 9.7 per 100,000 people with the use of the sensitive and specific algorithms, respectively. The total numbers of cases that were estimated by using the sensitive and specific algorithms for 2017 were 5,152 and 3,273 cases, respectively. All of the different estimations of the total number of cases identified with lung cancer and differentiated by region, sex and age groups are shown in Table 2.

**Table 2.** Prevalence rates and number of lung cancer cases estimated for 2017

|  | Prevalence rates 2017 |  | Number of cases 2017 |  |
| --- | --- | --- | --- | --- |
|  | Sensitive A. | Specific A. | Sensitive A. | Specific A. |
| Global | 15.3* | 9.7* | 5152 | 3273 |
| Men |  |  |  |  |
| 20 – 44 | 0.9* | 0.6* | 80 | 55 |
| 45 – 64 | 9.2* | 6.7* | 851 | 619 |
| 65 or more | 15.6* | 10.2* | 1444 | 947 |
| Women |  |  |  |  |
| 20 – 44 | 2.9* | 1.5* | 266 | 136 |
| 45 – 64 | 10.9* | 7.2* | 1005 | 664 |
| 65 or more | 16.3* | 9.2* | 1505 | 850 |
| Region |  |  |  |  |
| Atlantic | 8.9* | 5.5* | 609 | 378 |
| Bogota | 17.1* | 10.9* | 1079 | 688 |
| Central | 22.6* | 14.7* | 1934 | 1254 |
| Oriental | 12.1* | 7.6* | 679 | 427 |
| Orinoquia/Amazonia | 4.0* | 1.5* | 31 | 11 |
| Pacific | 14.7* | 9.2* | 819 | 514 |

\*In cases per 100,000. A: Algorithm

The distribution of prevalence rates and the number of total cases of lung cancer were also estimated by differentiating between the two health insuring regimes. Global prevalence rates for the contributory regime with the use of the sensitive and specific algorithms were 19.2 and 12.4 per 100,000 affiliates, respectively. For the subsidized regime, global prevalence rates with the use of the sensitive and specific algorithms were 9.7 and 5.8, respectively. This result represents a difference in approximately double of the number of cases diagnosed with lung cancer in the contributory regime over the subsidized regime. Prevalence rates shown in Table 3 indicated that in most of the groups that were studied, there were a greater number of cases identified in the contributory regime than in the subsidized regime.

**Table 3.** Prevalence rates and total number of cases differentiated by health insurer regime for 2017

|  | Subsidized regime |  |  |  | Contributory regime |  |  |  |
| --- | --- | --- | --- | --- | --- | --- | --- | --- |
|  | Prevalence rate |  | Number of cases |  | Prevalence rate |  | Number of cases |  |
|  | Sensitive A. | Specific A. | Sensitive A. | Specific A. | Sensitive A. | Specific A. | Sensitive A. | Specific A. |
| Global | 9.7* | 5.8* | 1331 | 794 | 19.2* | 12.4* | 3821 | 2478 |
| Sex and age group |  |  |  |  |  |  |  |  |
| Men |  |  |  |  |  |  |  |  |
| 20 – 44 | 0.1* | 0.03* | 3 | 1 | 1.3* | 0.9* | 77 | 54 |
| 45 – 64 | 13.1* | 8.2* | 271 | 170 | 21.0* | 16.3* | 580 | 450 |
| 65 or more | 39.9* | 26.9* | 392 | 264 | 83.6* | 54.2* | 1052 | 682 |
| Women |  |  |  |  |  |  |  |  |
| 20 – 44 | 3.8* | 1.4* | 152 | 58 | 2.1* | 1.4* | 115 | 79 |
| 45 – 64 | 11.0* | 6.9* | 249 | 156 | 25.8* | 17.3* | 756 | 508 |
| 65 or more | 24.3* | 13.4* | 264 | 145 | 81.7* | 46.4* | 1241 | 705 |
| Region |  |  |  |  |  |  |  |  |
| Atlantic | 6.8* | 4.0* | 286 | 167 | 12.2* | 8.0* | 323 | 210 |
| Bogota | 9.6* | 5.9* | 74 | 45 | 18.2* | 11.6* | 1005 | 643 |
| Central | 15.2* | 9.4* | 482 | 297 | 27.0* | 17.8* | 1452 | 958 |
| Oriental | 9.0* | 5.3* | 215 | 127 | 14.4* | 9.4* | 464 | 300 |
| Orinoquia/Amazonia | 3.5* | 1.4* | 18 | 7 | 4.9* | 1.6* | 14 | 4 |
| Pacific | 9.6* | 5.6* | 256 | 151 | 19.5* | 12.6* | 563 | 363 |

\*In cases per 100,000. A: Algorithm

## Discussion

The aim of this study was to develop an electronic algorithm to identify lung cancer-prevalent patients over 20-years-old in Colombia by using official databases, as well as to estimate prevalence rates of lung cancer by age, sex and geographic region. Two out of sixteen algorithms were selected as they had the closest prevalence rates to those rates reported by aggregated official sources such as The Global Observatory of Cancer, NCI and CAC. These algorithms were defined for this study as a sensitive algorithm and a specific algorithm because of their characteristics for identifying lung cancer cases, as described above. Using these two algorithms, the global prevalence rates for the contributory regime via the sensitive and specific algorithms were 19.2 and 12.4 per 100,000 affiliates, respectively. In the case of the subsidized regime, global prevalence rates via the sensitive and specific algorithms were 9.7 and 5.8, respectively.

As has been previously explained, aggregated prevalence rates were reported by official sources such as The Global Observatory of Cancer, NCI and CAC. However, the estimation methods used by the first two institutions were based on data obtained from samples of a limited number of patients with lung cancer attended at various IPSs around the country, thus introducing possible bias to the calculated rates [23,24]. Furthermore, prevalence rates reported by CAC were based on data provided by EPS; although it included information from 98% of the total population affiliated with the health system, these reported data could have underestimated the real prevalence, as it depended on passive reporting [25,26]. In the case of our research, the identification process involved the use of algorithms that take information from the usage of the reported services. This represents an advantage in comparison to the other sources, as our estimates rely on service usage rather than an active process of reporting or an estimation performed over a sample.

In this study, the estimations obtained by the two algorithms offer some additional advantages for policy-makers and clinicians. First, it allowed us to calculate prevalence rates differentiating between gender, regions and age groups. Second, it considered almost the total population affiliated with the contributory regime and the reported services that were utilized in the health system. Third, these estimations relied on the UPC administrative database, which represented consistent and validated data over the system, as it contains information with economical purposes. Fourth, it is possible to actively identify cases with lung cancer over the entire of population of patients attending to different providers. Finally, it is paramount to point out that these methods for identifying and estimating prevalence rates could help other LMICs to optimize their epidemiological surveillance system and to understand its impact in a context with different characteristics, compared to the developed world [27–30].

As LMIC countries enact changes to the population aging pattern, there is also a change in the burden of disease patterns from communicable to noncommunicable diseases (such as lung cancer), and efficient estimations are required to follow up on trends in public health [29,31–33]. Authors such as Sierra et al. [34] have conducted research to address this problem and collected information from thirteen different countries in Central and South America. They found that lung cancer ranged from first to third regarding causes in incidence rates over these countries, thus showing concordance with a change in the aging patterns. In 2018, a study performed by Pilleron et al. [35] estimated that there were 679,000 new cases of cancer in Latin America and the Caribbean, and the most frequent variants were prostate, colorectal and lung cancer. The findings in this study showed a global prevalence ranging between 9.7 and 15.36 cases per 100,000 that positions the disease in the top ten list of prevalent cancer in the country and that corresponds to the prevalence that was identified in our study.

Another important aspect of the lung cancer prevalence rates is inequality and differences in its distribution. It has been observed that developed countries have increased prevalence rates in comparison to developing countries [36]. There is also evidence showing that at the country scale, there is a trend of higher prevalence rates within lower socioeconomic groups that show differences in smoking rates or other exposure factors, such as chemical exposure, mining activities or higher exposure to air pollution [37–40]. For this study, the results showed two important patterns related to his idea. The first result showed that there is a higher prevalence rate of the disease in regions with highly polluted cities or concentrations of mining activities. The second trend found that prevalence rates that were calculated in the subsidized regime were lower than in the contributory regime. This finding differs from the literature, but it could be explained because of the inequalities in health care access that lead to a lower proportion of people being detected and treated with lung cancer. This last idea requires more research in the future.

This study had several strengths and limitations. As a strength, this research used real-world evidence from administrative claim databases that were considered to be highly reliable regarding its quality and validation procedure. Information calculated via the sensitive and specific algorithms showed results that were similar to those reported by other institutions and a range of possible prevalence rates, rather than an absolute number. In contrast, there were some concerns regarding the estimation of prevalence rates in the subsidized regime because these data were calculated based on information provided by the largest insurer in the capital city; however, this is the best information available, and it could not be easily extrapolated to the country level. There is a natural challenge in estimating real prevalence rates of lung cancer via the selected algorithms because they identify only patients with the disease and are detected by the system. However, there is a proportion of patients with the disease that are not detected by the system; hence, they were not included in our estimation. When taking into account that health coverage in Colombia reaches 99% of the total population [13], we assume that this difference was very small. Finally, it is also possible that information bias affected the study; however, quality and validation processes that were implemented by MoH helped to control this risk.

## Conclusion

Selected algorithms showed similar prevalence estimations to those reported by official sources and allowed us to estimate prevalence rates in specific aging, regional and gender groups for Colombia by using national claims databases. These findings could be useful to formulate new public health strategies and to measure prevalence trends in lung cancer at a national scale by using administrative databases.

## Data Availability

Data cannot be shared publicly because of an confidentiality agreement between Universidad Nacional de Colombia and the Ministry of Health of Colombia. Data are available from the integrated information system of the Ministry of Health of Colombia (https://www.sispro.gov.co/Pages/Home.aspx) for researchers who meet the criteria for access to confidential data

## Acknowledgments

The authors thank Juan Camilo Forero and Juan Sebastian Castillo for their commentaries and support during the research process.

